# Effectiveness of HIRARC in OSH Risk Management

**DOI:** 10.1101/2025.06.11.25329443

**Authors:** Wilson Lee Wui Siang, Mohd Rafee bin Baharudin, Anita binti Abdul Rahman, Nor Halim bin Hasan

## Abstract

Occupational safety and health (OSH) risk management is a systematic tool that assists organizations in identifying, assessing, and controlling work-related hazards to ensure the health, safety, and well-being of workers. In this regard, the Department of Occupational Safety and Health (DOSH) Malaysia has introduced Guidelines for Hazard Identification, Risk Assessment, and Risk Control (HIRARC) 2008 to assist structured hazard management. Hence, this study quantitatively assesses the impact of HIRARC implementation on OSH performance, focusing on its influence in improving workplace OSH risk management as well as reducing workplace accidents, dangerous occurrences, and health-related illnesses. Utilizing a sample of 390 DOSH-registered OSH practitioners across Malaysia, this study employs correlation and logistic regression analyses to quantify HIRARC’s effectiveness and identify key influencing factors. Findings show that effective HIRARC implementation correlates strongly with improved OSH risk management and significantly reduces workplace accidents as well as illnesses. These results underscore the importance of organizational participation, resource availability, and technical guidance in enhancing HIRARC’s impact. The study advocates broader HIRARC adoption across all sectors to improve workplace safety culture and create a better workplace environment.

## 1. Introduction

The implementation of occupational safety and health (OSH) risk management is vital in preventing workplace accidents and managing work-related hazards as it enables an organization to evaluate the risk of the hazard and take necessary measures to mitigate the risks systematically [1]. On top of reducing losses, costs and wastage of social resources, it is reported that having an effective risk management contributes towards improved operational performance, competitive value and reputation of an organization [2,3].

In Malaysia, there were 34,216 workplace accidents and 317 fatalities reported in year 2022. This translates into approximately 94 accidents and 1 fatality occurrence in a day [4]. As part of the initiative to overcome this issue, the Department of Occupational Safety and Health (DOSH) being the regulatory body for OSH in Malaysia, has introduced Guidelines for Hazard Identification, Risk Assessment and Risk Control (HIRARC) 2008 which aims to help Malaysian industries in managing workplace hazards [5].

The concept of risk management in Guidelines for HIRARC 2008 is based on a systematic approach that combines the process of recognizing, assessing, and measuring the risks, and finally reviewing the outcome [2]. This guideline is purported to be applicable for all sizes and types of sectors covered in Occupational Safety and Health Act (OSHA). Guidelines for HIRARC 2008 shares a similar conceptual approach with ISO 45001, an international standard for Occupational Safety and Health Management developed by International Organization for Standardization [6].

While several scholars have reported that conducting HIRARC is deemed to be simple, practical and commonly used [7,8], others have argued that there were instances where organizations still failed to identify all hazards within their workplace and there were also cases where the risk levels were wrongly classified leading to less effective control measures being proposed [9,10].

Though the guideline has been introduced for the past decade, there were little studies to gauge its effectiveness in managing OSH risk in Malaysia. Furthermore, the importance of OSH risk management has yet to be recognized by the industry especially in SMEs. OSH risk management allows organization to manage their workplace hazard systematically and assisting them to make necessary decision to address the risk. A search on the online database revealed that most of the present studies on the effectiveness of HIRARC were focused on measuring the implementation of HIRARC through case studies [7–9,11–13]. There were little studies conducted to measure the factors that influence the effectiveness of HIRARC in improving workplace safety and health. Furthermore, the studies were only focused on specific sector or size. Hence, this study would help the industry to understand the benefits of HIRARC and encourage them to implement risk management in their organization.

### 1.1. Methodology and Process-flow of Guidelines for HIRARC 2008

Guidelines for HIRARC 2008 was introduced by DOSH as part of the strategy to help employers to safeguard the health and safety of their workers and to reduce the numbers of occupational accidents, dangerous occurrences and occupational poisoning and diseases in Malaysia [14,15]. By implementing HIRARC, employers were able to identify all possible factors that may cause harm to the workers, evaluate the risk and plan for preventive measures to ensure that the risks are adequately controlled. There are four simple steps of conducting HIRARC, beginning with classifying work activities, followed by identifying workplace hazards, then conducting risk assessment, and finally deciding if control measures is required [16]. Guidelines for HIRARC 2008 adapts a semi-quantitative risk assessment approach using a five times five risk matrix that measures the severity and likelihood of the hazard. There are five levels respectively for severity and likelihood to describe the magnitude of the hazard whereby the descriptions were indicated using a holistic rubric. As a result, the risk is categorized into three levels, namely, low, medium, and high. After assigning the risk level, the following step would be determining the appropriate control measures. Guidelines for HIRARC 2008 conforms to hierarchy of control principles.

### 1.2. Factors that influence the effectiveness of HIRARC in improving workplace OSH risk management

Based on previous research, we have identified three main factors that influence the effectiveness of HIRARC in improving workplace safety and health, and the theories underpinning for these factors. Previous research discovered that organizational participation, organizational resources and providing technical guidance to conduct HIRARC were the main factors that affect the effectiveness of implementing risk management in an organization.

These factors were closely affiliated with Heinrich’s Domino Theory of accident prevention and Wernerfelt’s Resource-Based Theory. According to Heinrich [17], an accident would happen when the five conditions were satisfied in a sequence. HIRARC is considered the intervention of the third condition in domino series to remove unsafe act or physical hazards [18,19]. Additionally, resource-based theory developed by Birger Wernerfelt has been widely adapted in OSH studies to determine the critical elements of organizational resources towards safety performance [2,20,21].

#### 1.2.1. Organizational Participation

Studies have found that a successful OSH risk management depends significantly on the organizational participation that involves the commitment and participation of both the employers and workers. As a result, it was crucial that employers understand the importance of implementing HIRARC and were committed to implement the control measures that were suggested according to risk assessment. In this regard, employer can set priorities in managing OSH risk, better planning on engaging high risk activities, provide necessary allocation on OSH related matters, monitor the organization’s safety performance, and give incentives [8,9,15,22,23]. The involvement of workers through safety and health committee that consists of safety personnels, management representatives, employee representatives and other stakeholders were equally of importance in implementing HIRARC. Their knowledge in HIRARC would help the organization to communicate information regarding the hazard, control measures, safety instructions and other OSH related matters to be disseminated among the workers [5,9,22].

#### 1.2.2. Organizational Resources

Previous studies have identified that organizational resources that consist of financial resources, training on HIRARC and appointing designated personnel to conduct risk assessment would significantly affect the implementation of HIRARC in an organization. According to S. N. Ismail & Ramli [23], Kauthar A Rhaffor [11] and Marzuki et al. [5], small medium sized organizations (SME) have difficulty to implement risk control measures as they are struggling with financial constraints. Effective risk control measures often require installation of safety features or modification of process or equipment that involves additional financial allocation which the SMEs were reluctant to invest [11,14,22].

Studies have also discovered that employers, top management, and workers that were directly involved with risk assessment often lacks knowledge and not well-trained especially in SMEs. As a result, hazards were not thoroughly identified [8,9,15]. Furthermore, Liandar et al. [7] has reported that there were inconsistency of risk level rating being assigned between different projects with similar hazard and work activity in an organization due to safety knowledge barrier among the workers that conducted the risk assessment. Marzuki et al. [5] has described that employers typically in SMEs were reluctant to send their workers for HIRARC training as employment turnover rate was high and they were hard to be maintained. Hence, without proper trainings, the workers would need to self-interpret the terms and process of HIRARC that would lead to inaccurate assessment.

Another major constraint in effective implementation of HIRARC was lack of manpower to conduct risk assessment. Often SME organizations have difficulty to allocate staff to implement OSH related matters and the appointed personnel had to multi-task along with other job roles. Being occupied with other work obligations, the risk assessment were not done accordingly and was only prepared for documentation purposes [2,11,15,24]. Studies have shown that organization that having dedicated personnel well-equipped with OSH knowledge would be able to execute HIRARC effectively [11,15,25,26].

#### 1.2.3. Technical Guidance

For HIRARC to be implemented correctly, the guideline needs to be well-established, provide accurate, practical, and user-friendly guidance so that the users were able to understand the terms, process, and method of conducting risk assessment. In a study by Fathullah et al. [27], the author has conducted risk assessment on five case studies of construction and manufacturing accidents in Malaysia by adapting Guidelines for HIRARC 2008.

It was concluded that the guideline was able to identify the associated hazard and practical control measures were proposed based on the risk level priority. In another study the implementation of Guidelines for HIRARC 2008 in hydroelectric power generation plant has indicated that HIRARC was able to provide a systematic primary and secondary hazard analysis, classifying risk level and recommending control measures that would prevent accidents [28]. Furthermore, Pramudya [29] has reported that HIRARC is more simple to be conducted, less time consuming, and the control measures can be implemented immediately. These findings were consistent with studies conducted by Chia Kuang et al. [18], Hassan et. al [30], S.N. Ismail and Ramli et al. [22], and Kadir et al. [12] that HIRARC can be adapted at various sectors which include ports, education facilities, and mining. On the other hand, Marzuki et al. [15] has stressed that SMEs have difficulty to understand the terms and methodology to conduct HIRARC and the assessment were inaccurate as they only rely on self-learning and own interpretation.

### 1.3. Measuring safety performance of OSH Risk Management

The effectiveness of Guidelines for HIRARC 2008 in improving workplace safety performance for OSH risk management can be gauged using leading and lagging indicators [13]. Previous studies have shown that leading indicators were measured based on the ability of HIRARC to identify all possible workplace hazards and the practicality as well as effectiveness of control measures that were implemented [7–9,27,31,32]. On the other hand, lagging indicator is measured in accordance to the reduction of workplace accidents, dangerous occurrences, and occupational poisoning and diseases. Studies found that implementing HIRARC was able to reduce the level of risk of a hazard, and reduce workplace accidents when control measures were applied correctly [9,18,27,33].

## 2. Methods

### 2.1. Research Design and Instrumentation

In this cross-sectional study, data were collected using a survey questionnaire that consist of two main sections (1) Section 1: Socio Demographic of the respondents and (2) Effectiveness of risk management by adopting Guidelines for HIRARC 2008. There were twelve questions in Section 1 and twenty-six questions in Section 2. The questionnaires in Section 2 were developed based on the literature review, as shown in Table 1, and they are measured using a 5-point Likert scale (1 = Strongly Disagree, 5 = Strongly Agree). The instrument was subjected to a content validity evaluation and ten panellists were invited to review the instrument [34]. The panels consist of academicians, regulatory officers, and industrial practitioners. The content validity index (CVI) and content validity ratio (CVR) were above the minimum threshold as recommended by Gilbert and Prion [35], Lawshe [36] and Zamanzadeh et al. [24]. A pilot study was conducted to measure reliability of the questionnaire to provide consistency of the results. In this evaluation, the Cronbach’s Alpha coefficient was assessed and yields a reading of 0.912. Hence, this indicates high internal consistency and good respondent consistency.

**Table 1.**
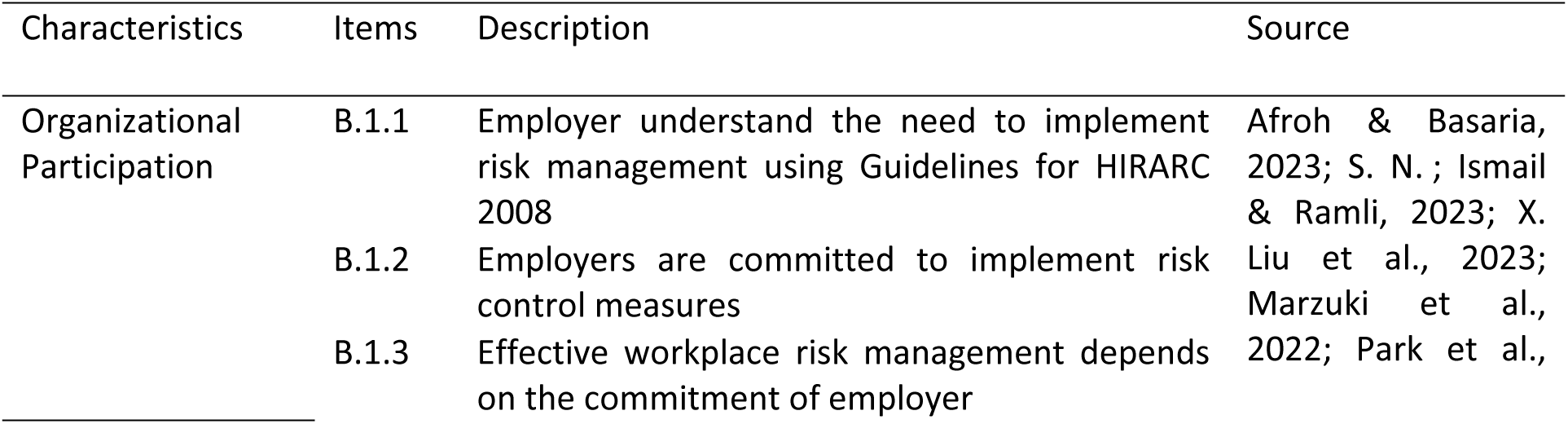
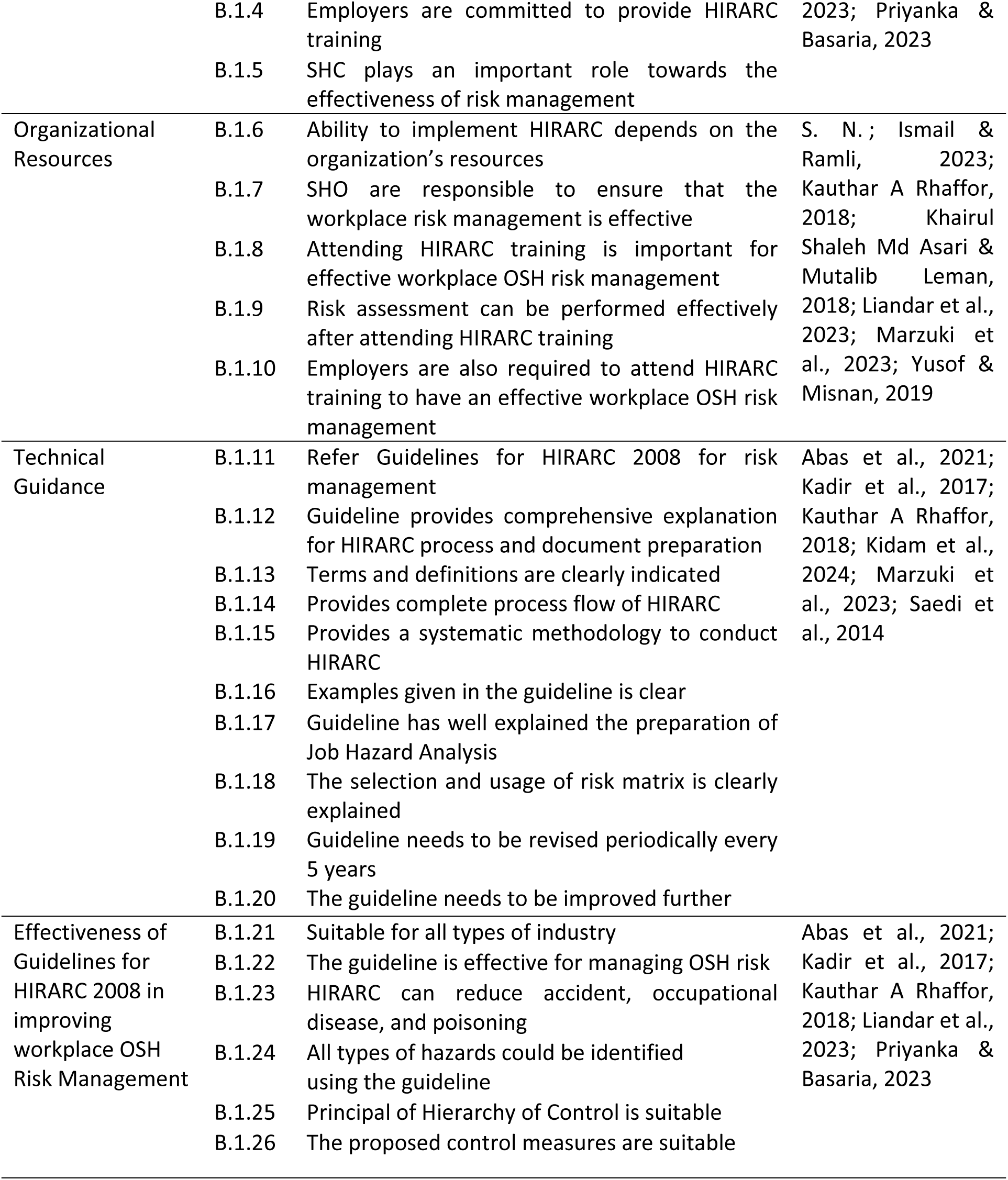
List of indicators and factors to measure the effectiveness of Guidelines for HIRARC 2008 in improving workplace OSH risk management.

### 2.2. Population and Research Sampling

The population and samples for this study were focused on DOSH registered safety and health practitioners. The sampling method was based on stratified random sampling that involves DOSH registered OSH practitioners that were involved with workplace OSH risk management and has established a safety and health committee in their respective workplace. The total population of DOSH registered OSH practitioners were 134,243 persons that consists of eighteen types of competencies [39]. By referring to Krejcie and Morgan’s sample size calculation [40], the minimum samples required was 384.

The survey questionnaire was distributed both physically and via email to the respondents, accompanied by a cover letter explaining the purpose of the study and an informed consent form stating that participation is entirely voluntary and that all information provided will remain confidential. The data were collected between 1^st^ of August 2024 to 31^st^ December 2024 from six regions in Malaysia in which (1) central region covers Kuala Lumpur, Putrajaya, and Selangor, (2) northern region covers the state of Perlis, Kedah, Penang, and Perak, (3) southern region covers the state of Negeri Sembilan, Malacca, and Johor, (4) east coast region covers the state of Kelantan, Terengganu, and Pahang, (5) the state of Sabah and (6) Sarawak.

The response rate was 64.56% whereby a total of 604 DOSH registered OSH practitioners were approached and 390 respondents have completed and returned the questionnaires. Majority of the respondents were male (82.3%) compared to female (17.7%). Among the respondents, 87.7% of them were safety and health officers (SHO), followed by 7.7% were OSH-Coordinator, 2.3% were site safety supervisor (SSS), 0.8% were Occupational Health Doctor (OHD), 0.5% were Steam Engineer, 0.5% were Authorized Gas Tester and Entrant Supervisor (AGTES), 0.3% were Hygiene Technician and 0.3% were Ergonomic Trained Person (ETP). Furthermore, 19% are currently in managerial position, 17.7% are executives, 53.6% are SHO, 2.8% are appointed as supervisors, 4.9% are consultants, 0.8% are academicians, and 1.3% are currently unemployed. Majority of the respondents, approximately 27.4% had seven to ten years of working experience. Additionally, majority of the respondents approximately 35.1% had seven to ten years of experience in OSH. Approximately 37.7% of the organization they are working with were established more than thirty years, followed by 25.9% that were established between twenty to twenty-nine years, 19.9% were established between ten to nineteen years, and 17.3% were established less than ten years. Respondents working in large organizations made up of 60.5% while those in medium sized organization were 28.8% and 10.6% were currently employed in small sized organizations. Construction sector made up the most respondents with approximately 40.5% and followed by 38.2% from manufacturing sector. Approximately 34.1% of the respondents were from central region, 19.7% were from northern region, 17.9% were from southern region, 13.3% were from east coast region, 8.2% were from Sabah, and 6.7% were from Sarawak. The socio demographic of the respondents is shown in Table 2.

**Table 2.**
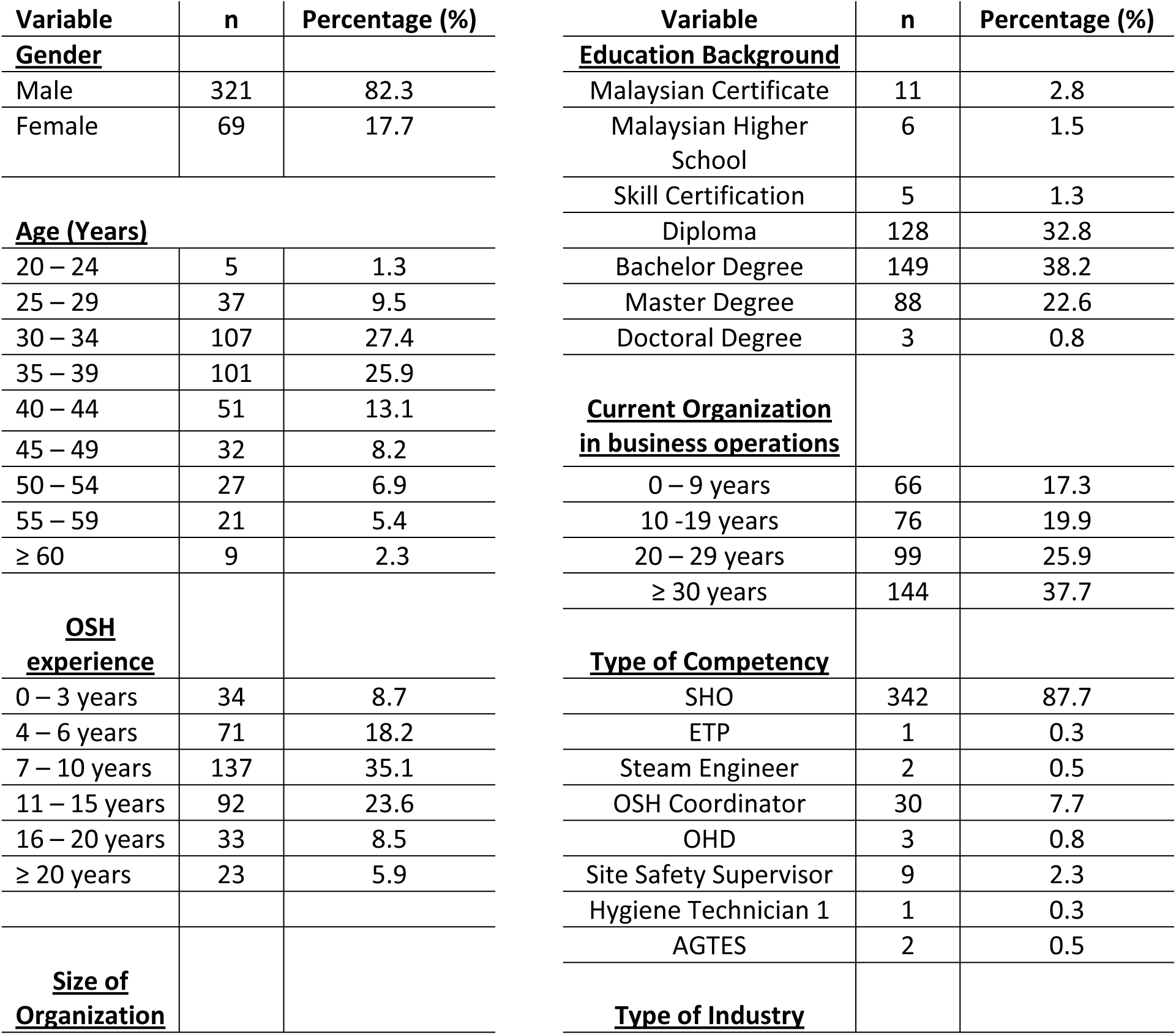
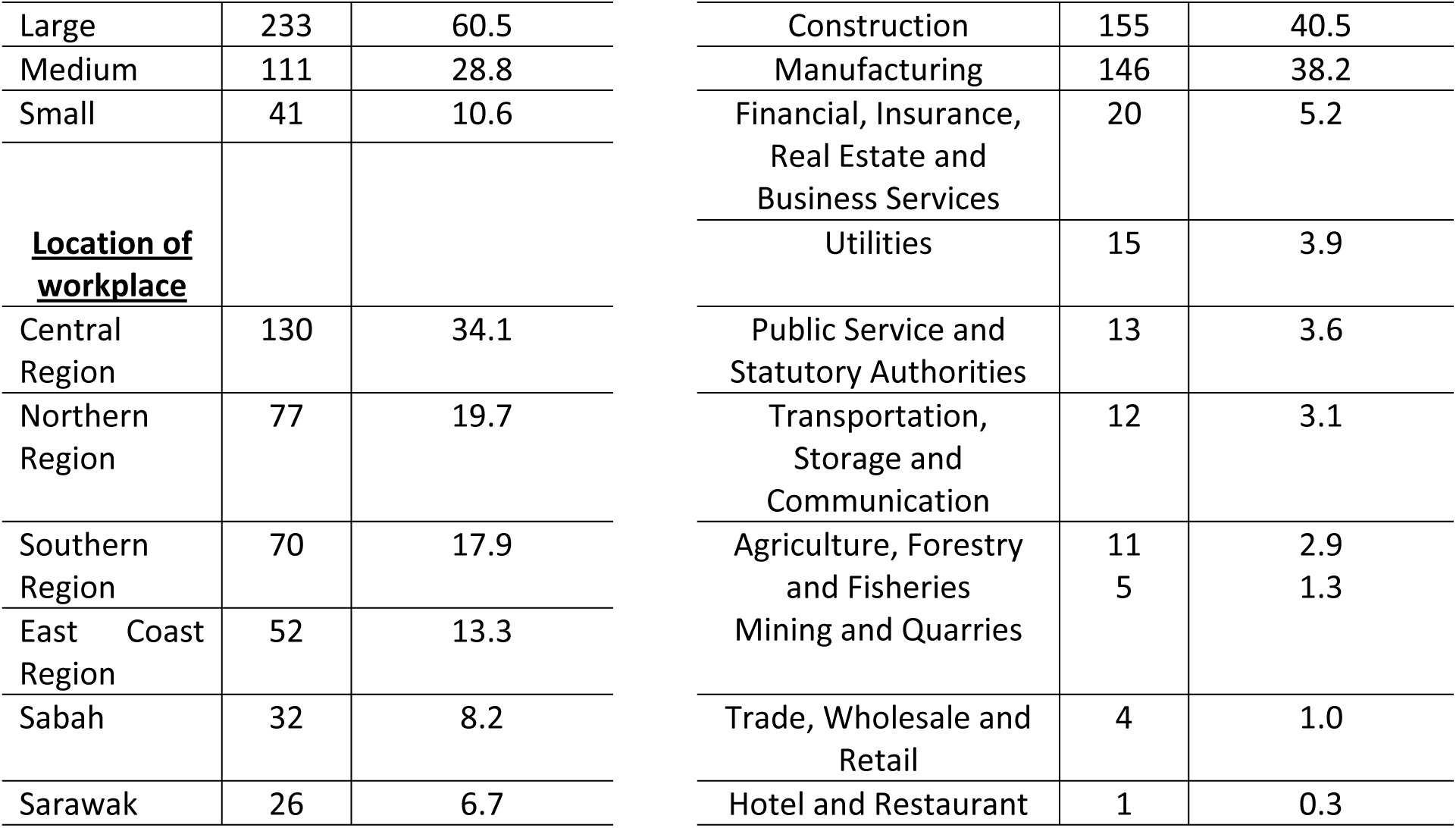
Socio demographic of respondents (n=390)

### 2.3. Data Analysis Method

Data obtained were analysed using Statistical Package for Social Science (SPSS) v25.0 to determine the correlation of implementing Guidelines for HIRARC 2008 towards improving workplace OSH risk management and reducing workplace accidents, dangerous occurrences, and occupational poisoning and diseases. A simple logistic regression was further performed to investigate the odds ratio of implementing Guidelines for HIRARC 2008 and reducing workplace accidents, dangerous occurrences, and occupational poisoning and diseases. The data were analysed based on a significant threshold of 0.05 (*p* = 0.05).

## 3. Results

There were two analysis conducted in this study where the first was determining the correlation of implementing Guidelines for HIRARC 2008 towards improving workplace OSH risk management and reducing workplace accidents, dangerous occurrences, and occupational poisoning and diseases, and secondly, performing a simple logistic regression to investigate the odds ratio of implementing Guidelines for HIRARC 2008 and reducing workplace accidents, dangerous occurrences, and occupational poisoning and diseases. A preliminary analysis was conducted to screen for outliers using the Mahalanobis distance (*D^2^*) assessment [41]. As a result, two multivariate outliers were identified and removed from the main data. The remaining three hundred and eighty-eight (388) data were further assessed using the normality test and the result as shown in Table 3 indicated that the data were not normally distributed.

**Table 3.** Normality test (Kolmogorov-Smirnov and Shapiro-Wilks Tests), n=388.

| Variable | Kolmogorov-Smirnov<sup>a</sup> |  |  | Shapiro-Wilks |  |  |
| --- | --- | --- | --- | --- | --- | --- |
|  | Statistic | df | Sig. | Statistic | df | Sig. |
| Implementatio<br>n | 0.086 | 388 | < 0.001 | 0.962 | 388 | < 0.001 |
| Indication | 0.132 | 388 | < 0.001 | 0.940 | 388 | < 0.001 |

### 3.1. Correlation Analysis

#### 3.1.1. Correlation between implementation of Guidelines for HIRARC 2008 and improvement to workplace OSH risk management

A non-parametric test was performed to determine the Spearman’s correlation coefficient on the implementation of Guidelines for HIRARC 2008 towards improving workplace OSH risk management and reducing workplace accidents, dangerous occurrences, and occupational poisoning and diseases. There were twenty items in the questionnaires (B.1.1 - B.1.20) that measures the implementation of Guidelines for HIRARC 2008 and there were five items (B.1.21-B.1.22 and B.1.24-B.1.26) to measure the improvement in workplace risk management.

The Spearman’s correlation coefficient showed in Table 4 showed that there was a strong positive correlation between implementation of Guidelines for HIRARC 2008 and improvement to workplace OSH risk management (*r_s_* = 0.820, p < 0.001).

**Table 4.** Correlation analysis on the total score in implementation of Guidelines for HIRARC 2008 and total score of improving workplace OSH risk management.

| Analysis | Numbers of respondents | Correlation Coefficient | Sig. (2 tailed) |
| --- | --- | --- | --- |
| Spearman's rho | 388 | 0.820 | < 0.001 |

#### 3.1.2. Correlation between implementing Guidelines for HIRARC 2008 and reducing workplace accidents, dangerous occurrences, and occupational poisoning and diseases

Additionally, the data were further analysed to determine the correlation between implementing Guidelines for HIRARC 2008 towards reducing workplace accidents. There was one item (B.1.23) to measure the implementation of Guidelines for HIRARC 2008 in reducing workplace accidents, dangerous occurrence, and occupational diseases. The Spearman’s correlation coefficient result is as shown in Table 5 indicate that there was a significant positive correlation between implementing Guidelines for HIRARC 2008 and reducing workplace accidents (*r_s_* = 0.671, p < 0.001).

**Table 5.** Correlation analysis on the total score in implementation of Guidelines for HIRARC 2008 and score of reducing workplace accidents, dangerous occurrence, and occupation diseases.

| Analysis | Numbers of respondents | Correlation Coefficient | Sig. (2 tailed) |
| --- | --- | --- | --- |
| Spearman's rho | 388 | 0.671 | < 0.001 |

### 3.2. Simple Logistic Regression

A simple logistic regression analysis was performed to determine the odds ratio of implementing Guidelines for HIRARC 2008 towards reducing occupational accidents, dangerous occurrences, and occupational poisoning and diseases. The analysis would be extended to include the three main factors that influence the implementation of HIRARC and their effect towards reducing occupational reducing occupational accidents, dangerous occurrences, and occupational poisoning and diseases. In the analysis, the respondents were categorized into two groups based on the median score where the first group having a total score of more than or equal the median scores, and second group than has a total score less than median score.

#### 3.2.1. Odds ratio of Organizational Participation towards reducing occupational accidents, dangerous occurrences, and occupational poisoning and diseases

There were five items in the questionnaire that measures organizational participation as a factor that influence the effectiveness of implementing HIRARC at workplace. The median score for this category was twenty-four. The logistic regression analysis found there was a positive significant relationship between respondents with higher total score of implementing organizational participation and reducing workplace accidents (*B* = 1.567, *SE* = 0.199, *Wald* = 62.120, *p* < 0.001). The estimated odds ratio as indicated in Table 6 showed that those who have higher score in organizational participation has 4.8 times to reduce workplace accidents, dangerous occurrences, and occupational poisoning and diseases (OR = 4.790, 95% CI = 3.245 – 7.072).

**Table 6.** Regression analysis between Organizational Participation, Organizational Resources, Technical Guidance, and Overall Implementation of HIRARC towards Reducing Workplace Accidents, Dangerous Occurrences, and Occupational Poisoning and Diseases.

| Item | Median Score | *B* | *SE* | *Wald* | *df* | *Sig.* | Crude OR | 95% CI |
| --- | --- | --- | --- | --- | --- | --- | --- | --- |
| Organizational Participation | 24 | 1.567 | 0.199 | 62.120 | 1 | <0.001 | 4.790 | 3.245, 7.072 |
| Organizational Resources | 21 | 1.640 | 0.204 | 64.714 | 1 | <0.001 | 5.157 | 3.458, 7.690 |
| Technical Guidance | 42 | 2.475 | 0.250 | 98.356 | 1 | <0.001 | 11.881 | 7.285, 19.377 |
| Overall Implementation | 87 | 2.348 | 0.240 | 96.131 | 1 | <0.001 | 10.468 | 6.546, 16.740 |

**Fig 1.**
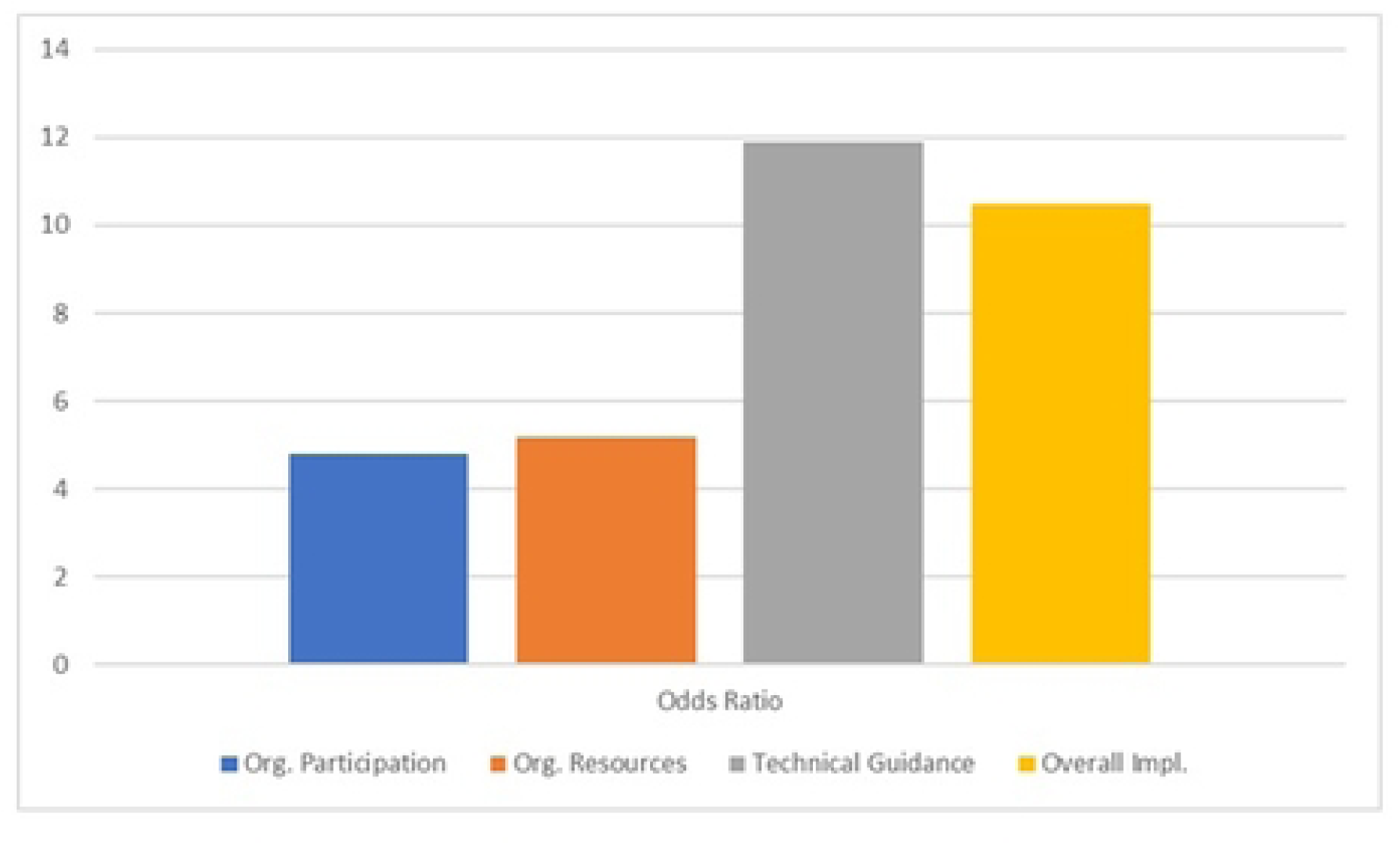
Comparison between logistic regression odds ratio for each key factors.

#### 3.2.2. Odds ratio of Organizational Resources towards reducing occupational accidents, dangerous occurrences, and occupational poisoning and diseases

In the questionnaire, there were five items pertaining to organizational resources in effective implementation of HIRARC. From the data collected, the median score for organizational resources was twenty-one. The logistic regression analysis also found that there was a positive significant relationship between the respondents with a higher total score of organizational resources and reducing workplace accidents (*B* = 1.640, *SE* = 0.204, *Wald* = 64.714, *p* < 0.001). Hence, the odds ratio indicated that allocating adequate resources to implement HIRARC has approximately 5.2 times to reduce workplace accidents, dangerous occurrences, and occupational poisoning and diseases (OR = 5.157, 95% CI = 3.458 – 7.690). The results of the simple regression analysis on organizational resources is shown in Table 6 below.

#### 3.2.3. Odds ratio of Technical Guidance towards reducing occupational accidents, dangerous occurrences, and occupational poisoning and diseases

A logistic regression analysis on the correlation between implementing risk assessment in accordance to the technical guidance provided in Guidelines for HIRARC 2008 and reducing workplace accidents, dangerous occurrences and occupational poisoning and diseases. There were ten items in the questionnaire with regards to technical guidance. Based on the data collect, the median score for technical guidance was forty-two. The logistic regression analysis found that there was a positive significant relationship between the respondents with a higher total score of technical guidance and reducing workplace accidents (*B* = 2.475, *SE* = 0.25, *Wald* = 98.356, *p* < 0.001). The analysis also indicates that respondents with a higher total score in following the technical guidance has approximately 11.9 times to reduce workplace accidents, dangerous occurrences, and occupational poisoning and diseases (OR = 11.881, 95% CI = 7.285 – 10.377).

#### 3.2.4. Odds ratio of Implementation of Guidelines for HIRARC 2008 towards reducing occupational accidents, dangerous occurrences, and occupational poisoning and diseases

Ultimately, a simple logistic regression analysis was conducted to determine the correlation between the overall implementation of Guidelines for HIRARC 2008 inclusive of organizational participation, and organizational resources towards reducing workplace accidents, dangerous occurrences and occupational poisoning and diseases. The median score for the overall implementation of Guidelines for HIRARC 2008 based on twenty items in the questionnaire was eighty-seven. The logistic regression analysis showed that there was a positive significant relationship between effective implementation of Guidelines for HIRARC 2008 and reducing workplace accidents (*B* = 2.348, *SE* = 0.24, *Wald* = 96.131, *p* < 0.001). The estimated odds ratio showed that implementing Guidelines for HIRARC 2008 effectively has approximately 10.5 times to reduce workplace accidents, dangerous occurrences, and occupational poisoning and diseases (OR = 10.468, 95% CI = 6.546 – 16.740).

## 4. Discussion

This study intends to discuss and analyse the relationship between effective implementation of HIRARC and improving workplace risk management as well as reducing occupational accidents, dangerous occurrences, and occupational poisoning and diseases. In this regard, this study comprises of three parts, firstly, to investigate the correlation between implementing HIRARC towards improving workplace OSH risk management. Secondly, to investigate the correlation between implementing HIRARC and reducing workplace accidents, dangerous occurrences and occupational poisoning and diseases. And thirdly, to determine the logistic regression of implementing HIRARC and reduction of occupational accidents, dangerous occurrences and occupational poisoning and diseases.

This study found that there was a strong positive correlation between effective implementation of HIRARC and improving workplace OSH risk management. Similarly, there was also a strong positive correlation between effective implementation of HIRARC and reduction of occupational accidents, dangerous occurrences and occupational poisoning and diseases. The effectiveness of implementing HIRARC was influenced by three factors, namely, organizational participation, organizational resources, and technical guidance of Guideline for HIRARC 2008. These findings were in line with previous studies by Afroh and Basaria [9], Bakri et al. [42], Buchari at al. [43], Chia Kuang et al. [18], and Othman et al. [31] where these researchers have concluded that HIRARC has helped organizations to identify workplace hazard and assisting them to make better decisions to mitigate the hazard according to the level of the risk. Furthermore, implementing HIRARC effectively were able to prevent workplace accidents, dangerous occurrence as well as occupational health related illness. This would create a conducive working environment, enhance productivity where workers feel more confident and potentially reduce absenteeism [27,43,44].

From a theoretical perspective, this study also indicates that effective implementation of HIRARC in accordance to Guidelines for HIRARC 2008 would significantly reduce workplace accidents, dangerous occurrences and occupational poisoning and diseases as much as 10.5 times. There were little studies that have measured the reduction of reduce workplace accidents, dangerous occurrences and occupational poisoning and diseases with regards to HIRARC implementation. Therefore, this study provides empirical evidence that allows organizations to understand importance of HIRARC and its magnitude of towards workplace accident reduction.

## 5. Conclusion

This study reveals that HIRARC is an effective tool in managing OSH risks and reducing workplace accidents as well as health-related illnesses. To maximize the impact, emphasis must be placed on dynamic participation of workers and employers towards OSH, allocating resource provisions, and systematic guideline application. The limitation of this study is that it only covers on DOSH registered practitioners. Thus, future research may include other OSH practitioners such as managers, business owners, and workers that are directly involved in the organization’s safety and health related matters. This would offer a comprehensive perspective towards the implementation of OSH risk management in an organization. Despite the use of simple logistic regression in this study, future research could benefit from multivariate or structural equation modelling (SEM) to refine the causal inference.

## Data Availability

Data cannot be shared publicly because the participants were informed of their confidentiality. Data are available from the Institutional Data Access from University Putra Malaysia for researchers who meet the criteria for access to confidential data.

## Acknowledgement

We appreciate the participation and consultation of members from Faculty of Medicine and Health Sciences, University Putra Malaysia in this study.

## Notes

### Competing Interest Statement

The authors have declared no competing interest.

### Funding Statement

The author(s) received no specific funding for this work.

### Author Declarations

This study protocol was registered and approved by Ethics Committee for Research Involving Human Subjects (JKEUPM), JKEUPM REF NO: JKEUPM-2024-1039

